# National research priorities for maternal and newborn health in The Gambia: a stakeholder-led prioritisation exercise

**DOI:** 10.64898/2026.07.09.26357708

**Authors:** Oghenebrume Wariri, Sainey Sanneh, Mamady Cham, Musa Marena, Oluwatosin Nkereuwem, Augusta Eneh, Lamin Makalo, Abdoulie Keita, Patrick Idoko, Haddy Tunkara Bah, Rachel Mendy, Baboucarr Cham, Phebian Ina Grant Sagnia, Alero Osas Ogebor, James Owolabi, Mustapha Bittaye, Momodou T. Nyassi, Bouba Manjang, Aduragbemi Banke-Thomas, Uduak Okomo

**Affiliations:** Vaccines and Immunity Theme, Medical Research Council Unit, The Gambia at London School of Hygiene and Tropical Medicine, Fajara, The Gambia; Department of Infectious Disease Epidemiology, London School of Hygiene and Tropical Medicine, London, United Kingdom; Ministry of Health, Banjul, The Gambia; Edward Francis Small Teaching Hospital, Banjul, The Gambia; School of Medical and Allied Health Sciences, University of The Gambia; School of Nursing and Midwifery, The Gambia; Centre for Maternal, Adolescent, Reproductive and Child Health, London School of Hygiene and Tropical Medicine, London, United Kingdom; Faculty of Epidemiology and Population Health, London School of Hygiene and Tropical Medicine, London, United Kingdom

## Abstract

Maternal and newborn mortality in The Gambia remains high despite expanded coverage of essential interventions, and progress towards SDG targets 3.1 and 3.2 has stalled. The National Reproductive, Maternal, Neonatal, Child and Adolescent Health (RMNCAH) Policy (2017 to 2026) ends in 2026, and a successor plan is in development. We aimed to identify and rank nationally defined maternal and newborn health (MNH) research priorities in The Gambia to inform the successor RMNCAH plan and align research investment with national health system needs.

We conducted a national, stakeholder-led MNH research prioritisation exercise in October 2023 using an adapted Child Health and Nutrition Research Initiative (CHNRI) method. Forty-six participants, including Ministry of Health policymakers and programme managers, clinicians, midwives, researchers, and development partners, took part in a two-day workshop. A starting set of 46 questions from the 2019 African Academy of Sciences (AAS) continental MNH prioritisation exercise and four questions from The Gambia’s National Health Research Agenda was expanded through facilitated discussion to a final list of 61 questions, organised by MNH grand challenge area and CHNRI research domain. Participants independently scored each question against four weighted criteria, and national rankings were compared descriptively with AAS continental and West African subregional rankings.

The priority list was dominated by delivery-focused research (49 of 61 questions, 80 percent), concentrated in better care during pregnancy and better postnatal care. The five highest-ranked priorities addressed management of obstetric emergencies before referral, retention and equitable distribution of the MNH workforce, neonatal resuscitation at peripheral facilities, maternal recognition of danger signs, and kangaroo mother care. Pre-transfer emergency obstetric management was the top national priority but was not prominent in AAS continental or West African subregional rankings.

This first national MNH research priority-setting exercise in The Gambia identifies a coherent set of implementation and health systems research priorities and surfaces context-specific questions, particularly pre-transfer emergency management, that were under-emphasised in continental rankings. The agenda provides an evidence base for the successor RMNCAH plan and for partner alignment in The Gambia and comparable high-burden settings.

## Introduction

Maternal and newborn health (MNH) is a central domain of population health, and a defining marker of how well health systems deliver essential care for women and their babies [1]. In The Gambia, recent expansions in coverage of antenatal, intrapartum, and postnatal interventions have not been matched by commensurate reductions in mortality [2–5]. The Gambia Demographic and Health Survey 2013 reported a maternal mortality ratio of 433 deaths per 100,000 live births and a neonatal mortality rate of 22 deaths per 1,000 live births [2]. By 2019 to 2020, the maternal mortality ratio had fallen to 289 per 100,000 live births, but the neonatal mortality rate had risen to 29 per 1,000 live births [5]. The 2023 UN Maternal Mortality Estimation Inter-Agency Group places, The Gambia among West African countries with persistently high maternal mortality, alongside neighbouring Senegal and Guinea-Bissau, highlighting both the scale of unfinished work and the heterogeneity of progress across the subregion [6]. Similarly, stillbirth remains an under-recognised dimension of this burden. The 2023 UN Inter-agency report on Stillbirths also estimates that sub-Saharan Africa accounted for around 48 per cent of global stillbirths in 2023, with intrapartum stillbirth concentrated in settings with limited access to skilled intrapartum care [7]. In The Gambia, recent facility- and population-based studies confirm that intrapartum stillbirth rates remain high and are strongly patterned by geographic access to emergency obstetric and newborn care [4, 8].

Most maternal and newborn deaths in The Gambia are preventable through known, low-cost interventions delivered through routine care [4, 8, 9]. Their persistence reflects bottlenecks in delivery, quality, referral, workforce capacity, and equity rather than an absence of effective technologies. Research that addresses these bottlenecks under real-world health-system conditions is therefore essential to identify what should be prioritised, how services can be strengthened, and where partner investment can have the greatest value [10]. However, in many low- and lower-middle-income countries such as The Gambia, the research that is funded and published is often shaped less by national health system priorities than by the interests of external funders and high-income institutions that lead global health partnerships [11–14]. This creates a mismatch between burden and agenda-setting power: countries with the highest MNH burden often contribute least to the research questions that shape global evidence, while locally relevant questions on implementation and health systems remain under-represented [11, 15].

Establishing explicit, nationally owned research priorities is one practical mechanism to address this imbalance, align research investment with country needs, and increase the likelihood that research informs policy and practice [16–18].

The Gambia’s National Health Policy identifies reproductive, maternal, newborn, child, and adolescent health (RMNCAH) as a strategic priority, while the 2021 National Health Research Agenda includes six ranked RMNCAH research questions, of which four are MNH-related [19, 20]. However, the limited number and breadth of these questions, and the absence of a structured priority-setting process focused specifically on MNH, mean they do not provide a sufficiently detailed national MNH research agenda to guide research investment, commissioning, and partner alignment.

At the continental level, the African Academy of Sciences (AAS) led a Child Health and Nutrition Research Initiative (CHNRI)-based MNH research prioritisation exercise in 2019, producing ranked priorities across Africa and identifying differences between Africa-based and international experts, particularly in delivery and health-systems research [21]. The exercise also showed meaningful variation across African subregions. Regional exercises can provide a useful foundation for national priority setting, but they cannot determine the relative importance of research questions within a specific country [18]. National health research agendas, therefore, need to be informed by priorities identified within the country, including those articulated by health workers and other stakeholders closest to service delivery.

With The Gambia’s National RMNCAH Policy, 2017 to 2026, [22] approaching the end of its implementation period and a successor plan in development, there is a timely opportunity for nationally ranked MNH research priorities to inform the next strategy and associated partner investments. This paper presents the findings of the first national research prioritisation exercise focused on maternal and newborn health in The Gambia [23, 24]. The aim was to identify and rank the MNH research priorities considered most important by national stakeholders and to examine how the resulting priorities relate to existing continental and subregional rankings. The exercise was intended to identify evidence gaps requiring policy and programme attention and provide a country-led basis for research commissioning, partner investment, and the research component of the successor RMNCAH strategy.

## Methods

### Study setting and design

The Gambia is a small West African country with a population of approximately 2.6 million. The public health system is structured into primary, secondary, and tertiary levels, with established referral pathways for obstetric and neonatal complications [19, 22]. The healthcare infrastructure includes one teaching hospital serving as the main referral centre, supported by a network of general and district hospitals, health centres, and clinics operated by communities, private organizations, and NGOs [25].

We conducted a national, stakeholder-led MNH research prioritisation exercise using an adapted CHNRI approach [26]. The exercise retained the core features of CHNRI: systematic development of research questions, predefined scoring criteria, independent stakeholder scoring, criterion weighting, and ranking of priorities [27]. The approach was adapted by using the existing continental MNH research priority list [21] and questions from the 2021 national research agenda [20] as the starting pool, followed by stakeholder review, contextualisation, and expansion. The exercise was reported in accordance with the REPRISE guideline for health research priority setting (S1 File) [28].

### Governance and participant recruitment

A Steering Committee convened by the Ministry of Health’s Directorate of Health Research defined the scope and national context of the exercise. The scope, including target population, geographic coverage, research domains, stakeholder groups, and policy context, is summarized in S2 File. The Steering Committee also agreed four scoring criteria in advance, informed by good-practice guidance for health research priority setting [18] and adapted to the Gambian context: potential to reduce disease burden; answerability and feasibility; potential impact on equity and vulnerable populations; and deliverability within the Gambian health system. The four scoring criteria, their component sub-questions, and the resulting criterion weights are shown in Table 1.

**Table 1:** CHNRI scoring criteria, sub-questions, and criterion weights.

| Criterion | Sub-questions | Weight |
| --- | --- | --- |
| Potential to reduce maternal and newborn disease burden in The Gambia | Are the findings likely to contribute to reducing maternal or newborn mortality in The Gambia? | 0.69 |
|  | Are the findings likely to contribute to reducing maternal or newborn morbidity in The Gambia? |  |
|  | Are the findings likely to contribute to reducing maternal or newborn disability or long-term adverse outcomes in The Gambia? |  |
| Answerability and feasibility in the Gambian context | Is the research question clearly framed, with outcomes or endpoints that can be defined and measured? | 1.39 |
|  | Is there sufficient technical, institutional, and human-resource capacity to conduct the research in The Gambia, or could this capacity reasonably be developed? |  |
|  | Is the research likely to obtain ethical and regulatory approval without major concerns? |  |
| Potential impact on policy, practice, and equity | Is the research likely to address an important evidence gap and contribute to improving maternal or newborn health policy, practice, or service delivery in The Gambia?<br>Are the findings likely to inform national planning, programme implementation, or resource allocation?<br>Is the research likely to benefit populations experiencing the greatest disadvantage, including people in lower-income households, marginalised groups, and those living in rural or hard-to-reach areas? | 0.89 |
| Deliverability within The Gambia's health system | Taking account of likely implementation requirements, including changes in practice, workforce capacity, supervision, referral systems, and transport infrastructure, could the findings be translated into policy or practice within The Gambia's health system?<br>Taking account of available resources, would the proposed intervention, strategy, or application of the findings be affordable and potentially cost-effective in The Gambia?<br>Is there sufficient government leadership, institutional capacity, and partner support to implement and sustain the proposed intervention, delivery strategy, or application of the findings? | 1.00 |

A two-day, in-person prioritisation workshop was held in October 2023 in Cape Point, The Gambia. Participants were purposively selected to represent the principal stakeholder groups involved in MNH policy, programme delivery, clinical care, research, and partner support.

Invitations were issued jointly by the Ministry of Health, which led government and policy engagement, and the MRC Unit The Gambia at LSHTM, which co-organised the exercise. Broader representation was sought through national clinical, academic, and research networks, including the national teaching hospital, the national university, and other research institutions. Participants did not receive financial incentives, although transport costs were reimbursed. The stakeholder groups invited included Ministry of Health policymakers and programme managers; frontline clinicians and implementers, including obstetricians, paediatricians, midwives, and nurses; researchers and technical experts; and development partners and donor representatives.

### Development and classification of the research question pool

The starting pool comprised 50 MNH research questions: 46 from the 2019 AAS continental CHNRI exercise [21] and four from The Gambia’s National Health Research Agenda [20]. The AAS questions provided a methodologically rigorous, expert-derived regional foundation for the national exercise, covering the continuum of maternal and newborn care and reflecting priorities relevant to sub-Saharan African settings. Using this established question set also allowed the Gambian rankings to be interpreted in relation to the continental and West African subregional priorities. The four MNH-related questions from the National Health Research Agenda were included to ensure that the starting pool reflected existing national priorities.

The starting questions were classified along two dimensions. The first was the MNH grand challenge area, which described the point in the continuum of care addressed by each question: better care during pregnancy; better care at birth; better postnatal care for women and their newborns; or better hospital care of sick newborns [29]. The second was the CHNRI research domain, which described the type of research required: description, discovery, development, or delivery [21]. Description questions addressed the burden or distribution of a problem; discovery questions addressed new biological, behavioural, or technical solutions; development questions addressed the design or testing of interventions; and delivery questions addressed the implementation or scale-up within real-world systems.

### Research question refinement and prioritisation

During the workshop, participants reviewed the starting questions through facilitated plenary and small-group discussions, considering their clarity, scope, and relevance to The Gambia. Broad questions were narrowed to make them more specific and answerable within the national policy and service delivery context, while overlapping questions were consolidated. Participants also generated additional questions informed by national epidemiology, health-system constraints, service delivery realities, and relevant MNH policy and strategy documents [9, 20, 22]. The consolidated question set was subsequently finalised for clarity and consistency, resulting in 61 questions spanning pregnancy, childbirth, and the early newborn period.

Participants first ranked the four scoring criteria from 4 (most important) to 1 (least important). They then independently scored all 61 research questions against the four criteria using a structured scoring sheet with three response options: yes, no, or informed but unsure. Independent scoring was used to minimise the influence of group dynamics and professional hierarchy.

### Analysis

The analytical approach followed that used in the AAS continental CHNRI exercise and included criterion weighting, calculation of weighted research priority scores, and ranking of questions.[21] Mean participant rankings were calculated for each scoring criterion and converted to criterion-specific weights; full details are provided in S3 File.

Responses to the question-scoring exercise were coded as 1 for yes, 0 for no, or 0.5 for informed but unsure. For each research question, responses were averaged across participants to produce a mean score for each criterion, ranging from 0 to 1. Each criterion-specific mean was multiplied by its corresponding weight. The weighted values were summed and divided by the sum of the criterion weights to produce a single weighted research priority score for each question, also ranging from 0 to 1. Higher scores indicated higher priority [30, 31].

Research questions were ranked from highest to lowest weighted research priority score to generate the national MNH research priority list. We described the distribution of questions by CHNRI research domain and MNH grand challenge area. We also compared the Gambian rankings descriptively with the continental and West African subregional rankings from the AAS CHNRI exercise.[21] Statistical comparisons were not undertaken because the exercises differed in scorer composition, weighting, and intended use. The descriptive comparison identified areas of alignment and divergence, as well as Gambian priorities for which no directly comparable question appeared in the continental or subregional rankings.

## Results

### Overview and distribution of prioritised research questions

A total of 61 MNH research questions were prioritised. Weighted research priority scores ranged from 0.92 for the highest-ranked question to 0.63 for the lowest-ranked question. The full ranked list is provided in S1 Table. Delivery-focused research accounted for 49 of the 61 questions, or approximately 80% of the priority list (Fig 1a). Development-focused questions formed a smaller proportion, while only a few questions were classified in the discovery or description domains.

**Figure 1.**
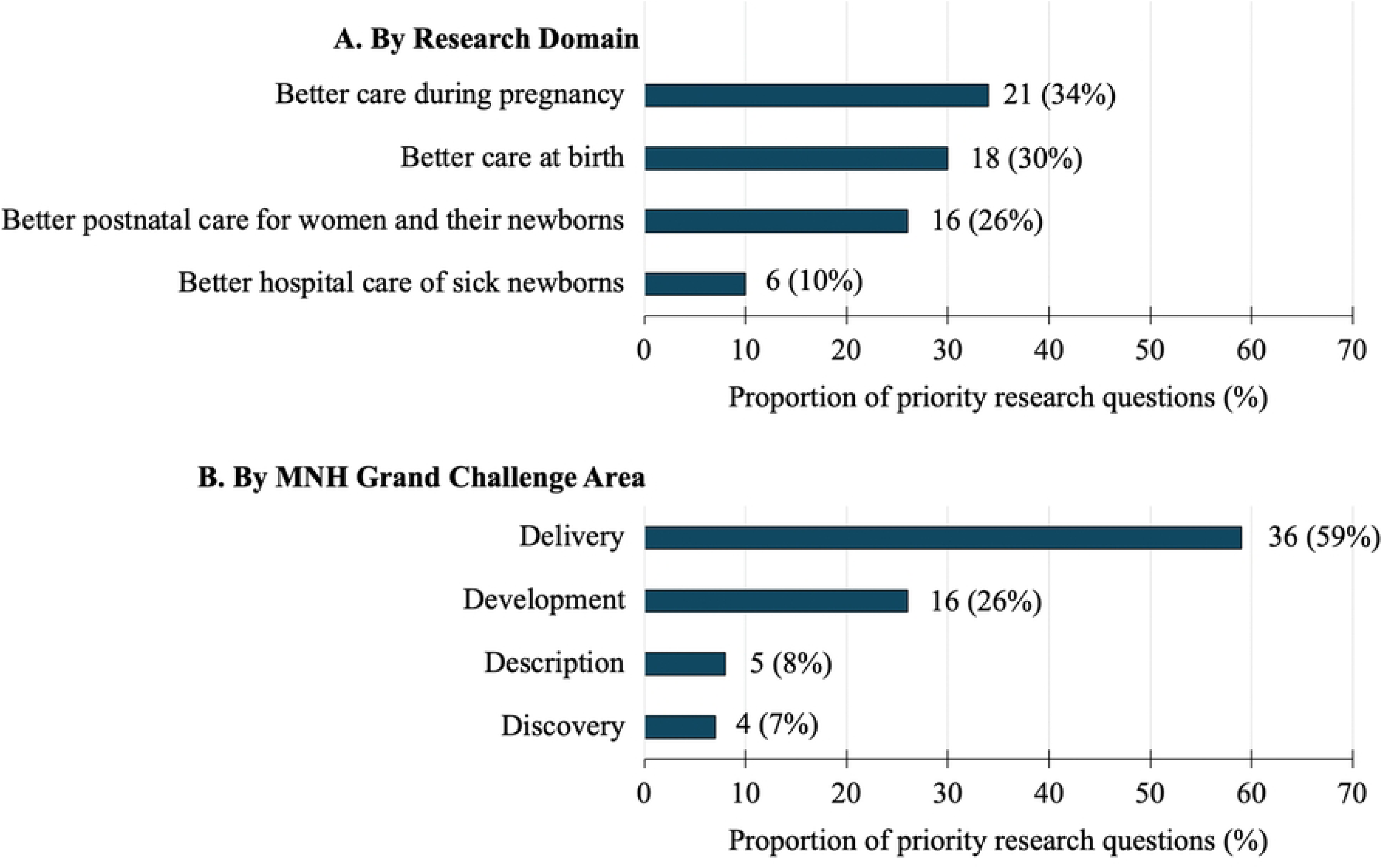
Distribution of the 61 prioritised maternal and newborn health research questions by (A) Research domain and (B) MNH grand challenge area. Bars show the proportion of all prioritised questions in each category.

Within the MNH grand challenge area, most questions focused on better care during pregnancy or better postnatal care for women and their newborns (Fig 1b). Fewer questions addressed better care at birth or hospital care for sick children.

### Top-ranked national priorities

Table 1 presents the ten highest-ranked national MNH research priorities, with research priority scores ranging from 0.84 to 0.92. The top-ranked question concerned strategies to improve the early management of obstetric emergencies at referring health facilities before transfer to the national referral hospital.

**Table 1.** Ten highest-ranked maternal and newborn health research priorities in The Gambia.

| Rank | Research Question | Question source | MNH grand challenge area | Research domain | Research priority score |
| --- | --- | --- | --- | --- | --- |
| 1 | What strategies can improve the early management and stabilisation of obstetric emergencies at referring health facilities before transfer to the national referral hospital? | Generated during national workshop | Better care during pregnancy | Delivery | 0.92 |
| 2 | What are the most effective strategies for attracting and retaining a trained health workforce? | Generated during national workshop | Better care during pregnancy | Delivery | 0.90 |
| 3 | What is the level of neonatal resuscitation competence among healthcare workers at peripheral hospitals, and how can it be improved? | Generated during national workshop | Better postnatal care for women and their newborns | Delivery | 0.89 |
| 4 | What is the effect of empowering mothers to recognise newborn danger signs on care-seeking and newborn health outcomes? | AAS CHNRI exercise | Better care during pregnancy | Delivery | 0.88 |
| 5 | What strategies are most effective in increasing community uptake of kangaroo mother care? | AAS CHNRI exercise | Better postnatal care for women and their newborns | Delivery | 0.87 |
| 6 | What is the prevalence of preterm birth in The Gambia, what factors are associated with it, and how can women at risk be identified and managed within the health system? | National Health Research Agenda | Better care during pregnancy | Development | 0.87 |
| 7 | What factors are associated with stillbirth in The Gambia, and how can high-risk pregnancies be identified, referred, and managed more effectively across levels of care? | National Health Research Agenda | Better care during pregnancy | Delivery | 0.86 |
| 8 | What is the effect of implementing the WHO postpartum haemorrhage bundle and related quality-of-care interventions on maternal outcomes in The Gambia? | Generated during national workshop | Better postnatal care for women and their newborns | Delivery | 0.85 |
| 9 | How ready is the health system to manage emergency obstetric complications, and what improvements are required? | AAS CHNRI exercise | Better care at Birth | Delivery | 0.85 |
| 10 | What strategies can improve the detection of hypertensive disorders of pregnancy at primary care level? | AAS CHNRI exercise | Better care during pregnancy | Development | 0.84 |

Initial question source indicates where each question entered the starting or expanded question pool. All questions were subsequently reviewed and refined through the national priority-setting process.

Other highly ranked questions addressed the attraction and retention of a trained health workforce, the evaluation of neonatal resuscitation skills among healthcare workers at peripheral hospitals, the effect of empowering mothers to recognise newborn danger signs, and the development and evaluation of strategies to increase community uptake of kangaroo mother care. The remaining priorities addressed the prevalence and determinants of preterm delivery and development of management protocols; identification, referral, and management of pregnancies at high risk of stillbirth; evaluation of the WHO postpartum haemorrhage bundle and quality-of-care interventions; assessment of health-system readiness for obstetric emergencies; and improved detection of pregnancy-induced hypertension at primary care level. Eight of the ten questions were classified in the delivery domain and two in the development domain.

### Comparison with continental and West African subregional priorities

Comparison of the Gambian rankings with the AAS continental and West African subregional rankings showed both areas of agreement and nationally specific priorities (Table 2). Several questions ranked highly across the exercises. Health workforce retention ranked second in The Gambia, third continentally, and seventh in West Africa; maternal recognition of newborn danger signs ranked fourth nationally and first in both AAS rankings; and detection of pregnancy-induced hypertension ranked tenth in The Gambia and second at both continental and subregional levels. Community uptake of kangaroo mother care also ranked highly in The Gambia and continentally, at fifth and sixth respectively, but was ranked twenty-second in West Africa. For four of the ten highest-ranked Gambian priorities, no directly comparable question was identified in the AAS rankings. These included the top-ranked national priority on early management of obstetric emergencies at referring facilities before transfer, as well as questions on preterm delivery, identification and management of pregnancies at risk of stillbirth, and implementation of the WHO postpartum haemorrhage bundle. Health-system readiness for obstetric emergencies ranked tenth in both The Gambia and West Africa, but nineteenth at the continental level.

**Table 2.** Comparison of nationally ranked maternal and newborn health research priorities in The Gambia with related continental and subregional priorities from the African Academy of Sciences–led CHNRI exercise.

| **Gambia Rank** | **Research Question** | **MNH Grand Challenges Category** | **Research Domain** | **AAS West African Rank<sup>a</sup>** | **AAS Continental Rank<sup>a</sup>** |
| --- | --- | --- | --- | --- | --- |
| 1 | What strategies can improve the early management and stabilisation of obstetric emergencies at referring health facilities before transfer to the national referral hospital? | Better Care during pregnancy | Delivery | Not applicable<sup>2</sup> | Not applicable<sup>2</sup> |
| 2 | What are the most effective strategies for attracting and retaining a trained health workforce? | Better Care during pregnancy | Delivery | 7 | 3 |
| 3 | What is the level of neonatal resuscitation competence among healthcare workers at peripheral hospitals, and how can it be improved? | Better postnatal care for women and their newborns | Delivery | Not applicable<sup>2</sup> | Not applicable<sup>2</sup> |
| 4 | What is the effect of empowering mothers to recognise newborn danger signs on care-seeking and newborn health outcomes? | Better Care during pregnancy | Delivery | 1 | 1 |
| 5 | What strategies are most effective in increasing community uptake of kangaroo mother care? | Better postnatal care for women and their newborns | Delivery | 22 | 6 |
| 6 | What is the prevalence of preterm birth in The Gambia, what factors are associated with it, and how can women at risk be identified and managed within the health system? | Better Care during pregnancy | Development | Not applicable<sup>2</sup> | Not applicable<sup>2</sup> |
| 7 | What factors are associated with stillbirth in The Gambia, and how can high-risk pregnancies be identified, referred, and managed more effectively across levels of care? | Better Care during pregnancy | Delivery | Not applicable<sup>2</sup> | Not applicable<sup>2</sup> |
| 8 | What is the effect of implementing the WHO postpartum haemorrhage bundle and related quality-of-care interventions on maternal outcomes in The Gambia? | Better postnatal care for women and their newborns | Delivery | Not applicable<sup>2</sup> | Not applicable<sup>2</sup> |
| 9 | How ready is the health system to manage emergency obstetric complications, and what improvements are required? | Better Care at Birth | Delivery | 10 | 19 |
| 10 | What strategies can improve the detection of hypertensive disorders of pregnancy at primary care level? | Better Care during pregnancy | Development | 2 | 2 |

## Discussion

### Summary of principal findings

This first national research prioritisation exercise, focused on maternal and newborn health in The Gambia, produced three main findings. First, stakeholders strongly prioritised delivery-focused research, with approximately four in five questions classified in the delivery domain. Second, priorities were concentrated on better care during pregnancy and better postnatal care for women and their newborns, while fewer questions focused on care at birth or on hospital care for sick children. Third, although several highly ranked Gambian priorities were also prominent in the continental and West African subregional rankings, the national exercise assigned greater importance to health system bottlenecks. The clearest example was early management of obstetric emergencies at referring facilities before transfer, which ranked first nationally but did not feature among the corresponding top continental or subregional priorities. Questions on health-system readiness for obstetric emergencies and on the implementation of the WHO postpartum haemorrhage bundle were also ranked higher in The Gambia. These differences show that continental and subregional exercises can identify shared research concerns, but national prioritisation is needed to determine which challenges are most urgent within a particular health system.

### Implementation over innovation: what stakeholders are saying

The central finding of this exercise was the strong emphasis on delivery-focused research. Gambian stakeholders, including Ministry of Health leaders, program managers, frontline clinicians, midwives, and those responsible for MNH service delivery, placed the greatest priority on questions concerned with how existing interventions could be implemented effectively within the health system. This does not imply that technological innovation or biomedical discovery is unimportant. Rather, it suggests that stakeholders regarded implementation, service delivery, and quality of care as the more immediate research priorities for reducing maternal and newborn mortality in The Gambia.

Three factors may have contributed to this pattern. First, the major causes of maternal and newborn mortality in The Gambia—including obstetric haemorrhage, hypertensive disorders, sepsis, birth asphyxia, and complications of prematurity—are conditions for which evidence-based interventions are available and reflected in national and international guidance. These include active management of the third stage of labour, magnesium sulphate for eclampsia, neonatal resuscitation, kangaroo mother care, and timely referral for emergency obstetric care [32]. The unresolved questions are therefore often about implementation: whether services are available, whether staff have the required skills and resources, and whether care is delivered consistently and at the required standard. Second, the weighting of the prioritisation criteria may have reinforced this orientation. Participants placed high importance on deliverability within the Gambian health system, as well as on answerability and feasibility. These criteria are likely to favour research questions that can inform action within existing system constraints. Third, the workshop’s composition may also have influenced the results. Most participants were directly involved in policy, programme management, or service delivery, and were therefore closely exposed to the operational constraints affecting maternal and newborn care. A priority-setting exercise involving a different balance of stakeholders, such as a larger proportion of basic scientists or research funders, might have produced a different distribution of research domains. The content of the prioritised questions was consistent with this emphasis. Stakeholders focused on retaining trained health workers, strengthening emergency referral systems, improving recognition of danger signs, assessing neonatal resuscitation skills, increasing uptake of kangaroo mother care, and evaluating implementation of standardised care bundles such as the WHO postpartum haemorrhage bundle. These are not peripheral operational issues. They are the conditions that determine whether evidence-based interventions are delivered effectively and whether they translate into reductions in preventable maternal and newborn deaths.

This stakeholder focus on delivery differs from global health research funding patterns, in which discovery, product development, and technological innovation often receive greater visibility and investment [13, 33]. For research funders, policymakers, and commissioners of MNH research in The Gambia, the clear implication is that the most relevant research for preventing mortality in the short term will likely stem from health systems, implementation, and quality improvement studies that target barriers in frontline and referral care, rather than from discovery-phase research alone [32, 34].

### Why national prioritisation matters despite continental rankings

Research priorities may be set at global, regional, national, or other levels, and their relevance will vary by context.[18] Some priorities may be shared across settings, while others will reflect epidemiological, health-system, and policy needs. Because the Gambian exercise was built on the AAS continental MNH CHNRI exercise, some overlap with continental and West African subregional priorities was therefore expected. Of the 50 starting questions, 46 originated from the AAS exercise [21], and several highly ranked Gambian priorities were also prominent in the continental or West African subregional rankings. This overlap reflects service-delivery and health-system challenges shared across African settings with high maternal and newborn mortality. The national exercise established which of these shared challenges were most important in The Gambia and identified additional bottlenecks arising from the country’s health system context.

The value of the Gambian exercise lay not only in simply producing different priorities, but in assigning national importance and order to the research questions. It converted a broad continental question pool into a ranked list shaped by Gambian policymakers, clinicians, programme staff, researchers, and partners. Participants also added questions grounded in national epidemiology, service delivery realities, health system structure, and policy needs. The clearest example was the top-ranked Gambian priority on improving the management of obstetric emergencies at referring facilities before transfer to the main referral hospital, highlighting national concerns about referral readiness, early stabilisation, and capacity at first-contact facilities.

### Implications for the successor RMNCAH plan and partner alignment

The Gambia’s National RMNCAH Policy (2017 to 2026) [22] is nearing the end of its implementation period, and a successor plan is under development. The priorities identified through this exercise correspond closely with major areas of the current policy, including quality of care, emergency obstetric and newborn services, postnatal care, referral systems, and health workforce development. They provide a clear basis for defining the research priorities of the successor plan and for linking those priorities to routine planning and budgeting processes, including the Medium-Term Expenditure Framework and annual operational plans.

The ranked list also offers a practical reference point for research commissioning and partner engagement. For development partners and research funders, closer alignment of funding calls, technical support, and reporting requirements with nationally defined MNH priorities could improve coherence across investments and increase the likelihood that research findings inform programme design, resource allocation, and service delivery in The Gambia. This is particularly important in the current global health funding environment. Growth in global health research funding has slowed, resources remain concentrated in high-income institutions, and implementation-oriented research continues to attract limited dedicated investment [35–37]. A transparent national priority list can therefore support accountability by enabling assessment of whether research commissioned, funded, and conducted in The Gambia reflects stated national needs and whether the resulting evidence is used in policy and practice [11, 33]. Even modest domestic investment in nationally defined priorities would further strengthen country ownership and support continuity of MNH research aligned with health-system needs [36, 38].

### Strengths and limitations

This exercise has several strengths. It engaged a broad cross-section of MNH stakeholders, including Ministry of Health leaders, programme managers, clinicians, midwives, researchers, and development partners, strengthening the legitimacy and policy relevance of the resulting priorities [26]. The use of a structured CHNRI-adapted approach, predefined criteria, independent scoring, and explicit criterion weighting made the value judgements underpinning the rankings transparent and reproducible. The process strengthened the relevance and interpretability of the findings. The AAS question pool allowed the Gambian priorities to be interpreted alongside continental and subregional rankings, while national agenda questions and stakeholder-generated questions ensured that the final list reflected Gambian policy priorities and service delivery realities.

Several limitations should be acknowledged. First, direct participation from communities, service users, and research funders was limited. The priorities therefore mainly reflect the perspectives of policymakers, implementers, clinicians, researchers, and partners who attended the workshop. Future exercises should more deliberately include women’s groups, civil society organisations, service-user representatives, and funders, in line with good-practice recommendations for health research priority setting [39]. Second, the starting question set drew heavily on the AAS continental exercise. This provided a rigorous foundation and enabled comparison with continental and subregional rankings, but it may also have shaped the range of questions considered. The addition of context-specific questions during the workshop partly mitigated this limitation.

## Conclusion

This national MNH research prioritisation exercise provides a practical foundation for strengthening the research component of The Gambia’s successor RMNCAH plan. The findings point to the need for research investment focused on implementation, quality of care, referral systems, workforce retention, and frontline service delivery, rather than treating MNH research priorities as primarily technology-led or discovery-driven. The exercise also reinforces the importance of national prioritisation, even where continental and subregional rankings exist.

Wider rankings can identify shared MNH concerns, but national processes are needed to determine which bottlenecks require urgent attention within a specific health system. The value of these priorities will depend on their uptake into the successor RMNCAH plan, research commissioning, partner planning, and funding decisions.

## Data Availability

The minimal dataset is available at Open Science Framework (OSF) via https://doi.org/10.17605/OSF.IO/VMCWY

## Acknowledgements

We thank the maternal and newborn health experts and stakeholders who participated in the research prioritisation exercise, the Ministry of Health of The Gambia for its leadership and engagement, and the staff of the Medical Research Council Unit The Gambia at LSHTM for logistical support.

## Supporting information

**S1 File. REPRISE (REporting guideline for PRIority SEtting of health research) checklist**.

**S2 File. Scope of the national maternal and newborn health research priority-setting exercise in The Gambia**

**S3 File. Calculation of criterion weights**

**S1 Table. Ranked maternal and newborn health research priorities in The Gambia, with research domains, MNH grand challenge areas, criterion-specific scores, and overall research priority scores**

